# Manifestations, Diagnosis and Treatment of Osseous Rosai-Dorfman Disease: A Systematic Review

**DOI:** 10.1101/2023.08.30.23294833

**Authors:** Abhimanyu Agarwal, Mostafa Abozeed

## Abstract

**BACKGROUND:** We hypothesize that lymphadenopathy and extraosseous manifestation affect the management protocol and our aim is to aid in development of standardized diagnosis and treatment protocols.

**METHODS:** PubMed and Google Scholar were searched from January 2012 through December 2022 using keywords “Rosai-Dorfman”, “Rosai and Dorfman,” and “bone“. Out of 98 researches, meta-analysis of 51 full-text studies involving 53 individuals with osseous RDD was conducted.

**RESULTS:** A significant association was found between lymphadenopathy with steroid therapy (OR: 20.125, 95%CI: 3.02, 134.09, P=0.0004). Extraosseous RDD showed significant association with steroid therapy (P=0.0145), T1-isointense MRI (OR: 0.173, 95%CI: 0.0304, 0.986, P=0.035), T1-hyperintense MRI (OR: 0.16, 95%CI: 0.0314, 0.815, P=0.019) and T2-hyperintense MRI (OR: 0.115, 95%CI= 0.0206, 0.6456, P=0.007).

**CONCLUSION:** Osseous RDD is rare and lacks standardized diagnosis and treatment protocols. Patients with lymphadenopathy were 9 times more likely to receive steroids, and those with extraosseous manifestations were less likely to undergo MRI. RDD is often not initially considered in differential diagnoses, leading to worsened conditions. We propose that further testing is required in affected individuals to develop standardized protocols.

**Key Points:**

1. Revealed variations in osseous and extraosseous involvement and immunohistochemistry played a crucial role in diagnosis.
2. Found association of lymphadenopathy and extraosseous involvement with steroid therapy and diagnostic modalities of RDD.
3. Limitations include reliance on case reports (low-quality evidence), potential inclusion bias in reported data.

## INTRODUCTION

Rosai-Dorfman disease (RDD) is a rare histiocytosis characterized by accumulation of S100-positive macrophages that commonly demonstrate emperipolesis. Historically regarded as an inflammatory/benign disorder, recent advancements have shown the presence of mitogen activated protein kinase (MAPK) pathway mutations (specifically KRAS, NRAS) underlying the pathogenesis in these disorders. Recently, this has been recognized as a distinct histiocytic neoplasm by the World Health Organization and International Consensus Classification of hematopoietic tumors^1,2^.

Per the Histiocyte Society classification, RDD is considered under the ‘R’ group of histiocytosis and comprises nodal, extra nodal and familial cases^3^. Classic (nodal) RDD mainly affects younger people, mostly African, with a minor male predominance. Comparatively, patients with Cutaneous RDD tend to be older, female, and more likely Asian or white^4^. Skin, nasal cavity, bone, orbital tissue and central nervous system (primarily dural) are common extranodal sites of involvement in classical RDD^5^. Lytic lesions with clearly visible sclerotic borders on radiographs are linked to nodal diseases that affect the bones^4^.

By morphology, RDD histiocytes are characterized by enlarged round to oval nuclei with distinct nucleoli, open chromatin and abundant pale eosinophilic cytoplasm with engulfment of mononuclear inflammatory cells (emperipolesis). By immunohistochemistry, RDD cells are positive for CD68, S100, OCT2 and cyclinD1; they are negative for langerhans cell markers (CD1a, langerin). RDD cells are frequently associated with dense lymphoplasmacytic infiltrates^6^. Although there is no standardized treatment and established protocol currently, systemic therapy is suggested for multifocal unresectable extranodal illness. Systemic therapies such as corticosteroids, sirolimus, radiation, chemotherapy and targeted immunomodulatory therapy can be used. For cases of unifocal extranodal disease or symptomatic airway, cranial, spinal or sinus disease, surgical excision may be required^7,8,9^.

## MATERIAL AND METHODS

### Search strategy

This systematic review and meta-analysis was conducted following the Preferred Reporting Items for Systematic Reviews and Meta-Analysis statement. PubMed, Scopus and Embase databases were systematically searched from 1 January 2012 until 31 December 2022. Investigators performed an independent literature search using the search terms ‘rosai dorfman disease’, ‘RDD’, ‘osseous involvement’, ‘extranodal involvement’ and ‘primary bone disease’. Institutional review board approval was not sought because of the use of publicly available data.

### Study selection

This systematic review and meta-analysis included all studies published from 1 January 2012 until 31 December 2022 in English-language only if they met the following eligibility criteria:

1. Observational studies, case reports, case series.
2. Patients with clinical diagnosis of Rosai Dorfman Disease primarily involving bone.
3. The site of bone involvement and presence or absence of lymph node involvement were among the clinical outcomes of interest.
4. For bone involvement assessments, we considered publications that discussed the results obtained using recognized tools like MRI, CT scan.
5. Any diagnosis of RDD or associated symptoms given using an established categorization system was considered.

Exclusion criteria:

1. Patients who had diseases that mimicked RDD.
2. Studies discussing pathophysiology of Rosai Dorfman Disease,
3. Did not present original data or were editorials systematic reviews/meta analysis.
4. We excluded articles that exclusively reported on substitute measurements.

All results were exported to Zotero, where duplicates were eliminated. Researchers independently reviewed and included publications by agreement, while a senior author arbitrated any disagreements.

### Data collection

Information from each study, including author names, publication year, country, and baseline variables, was gathered using a structured data collecting form and hazard risks (HR)/ Odd’s ratio (OR) with 95% CI.

### Data Synthesis and Analysis

We conducted a meta-analysis focused on exploring the potential associations of lymphadenopathy and extraosseous involvement with steroid therapy and the diagnostic modalities of RDD. By employing Microsoft Excel, the gathered data from multiple studies was organized, allowing for systematic comparison and aggregation. Utilizing statistical functions within Excel, we calculated OR, 95% confidence intervals, and conducted the chi-square test of independence to identify potential patterns or correlations between lymphadenopathy, extraosseous involvement, steroid therapy, and RDD diagnostic modalities. We also used Excel to generate graphical representations.

## RESULTS

The total number of cases reported was 51, with a total of 53 patients. The distribution of cases based on sex showed 18 were male, 34 were female, and 1 case did not have its sex reported. The mean age of the patients was 34.06 years with a standard deviation of 18.59 (age range 1-76 years). The average age for males was 28.94 ± 17.81 years, while for females it was 37.44 ± 18.42 years. Figure 1 shows the inclusion and exclusion of the studies included in this review.

**Fig. 1.**
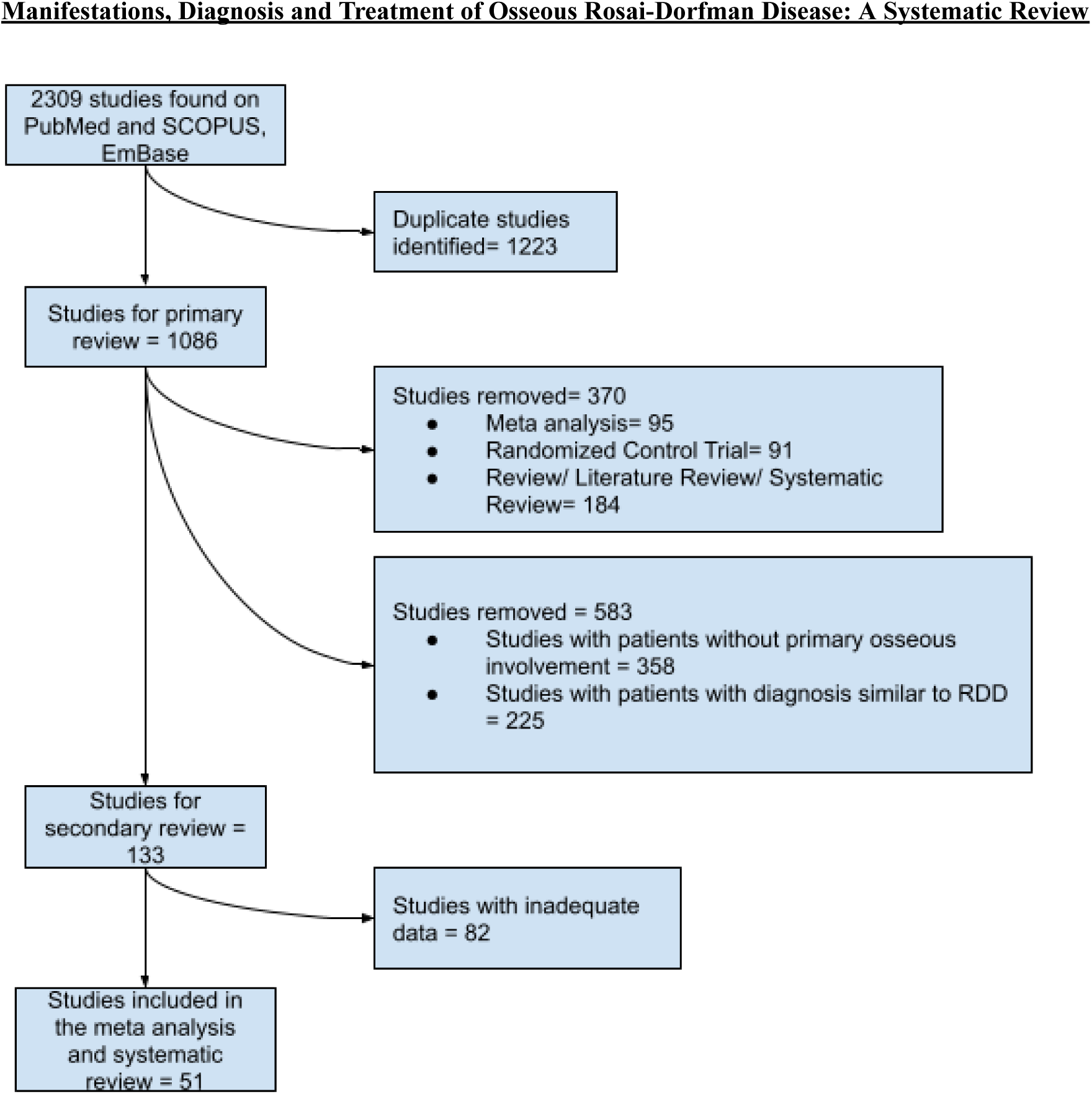
Study selection using PRISMA technique.

In terms of skeletal involvement, the following bones were affected: 9 cases involved the cranium, 9 involved the tibia, 8 involved the vertebra, 6 involved the femur, 3 involved the patella, 3 involved the radius, 2 involved the humerus, 2 involved the maxilla, and 1 case each involved the ilium, malleolus, mandible, metacarpal, nasal turbinate, pharynx, ribs, sacrum, ulna, sternum, and talus. Two cases did not report the specific bone affected.

Regarding lymphadenopathy, 25 patients (47.17%) showed no lymph node enlargement, 17 patients (32.08%) did not report lymph node involvement, and 11 patients (20.75%) had lymphadenopathy. The number of lymph nodes involved varied among the patients. Multiple lymph nodes were found in a total of 4 studies, involving 6^39^, 4^34^, 3^52^ and 2^54^ nodal sites while 7 studies^34,40,45,46,50,51,56^ reported only one lymph node involvement. The types of lymph nodes affected (Fig. 2) included 8 cases involving the cervical region, 3 involving the mediastinum, 3 involving the inguinal region, and 1 case each involving the parotid, presacral, iliac, sacral, intermedullary, axillary, and pelvic regions.

**Figure 2:**
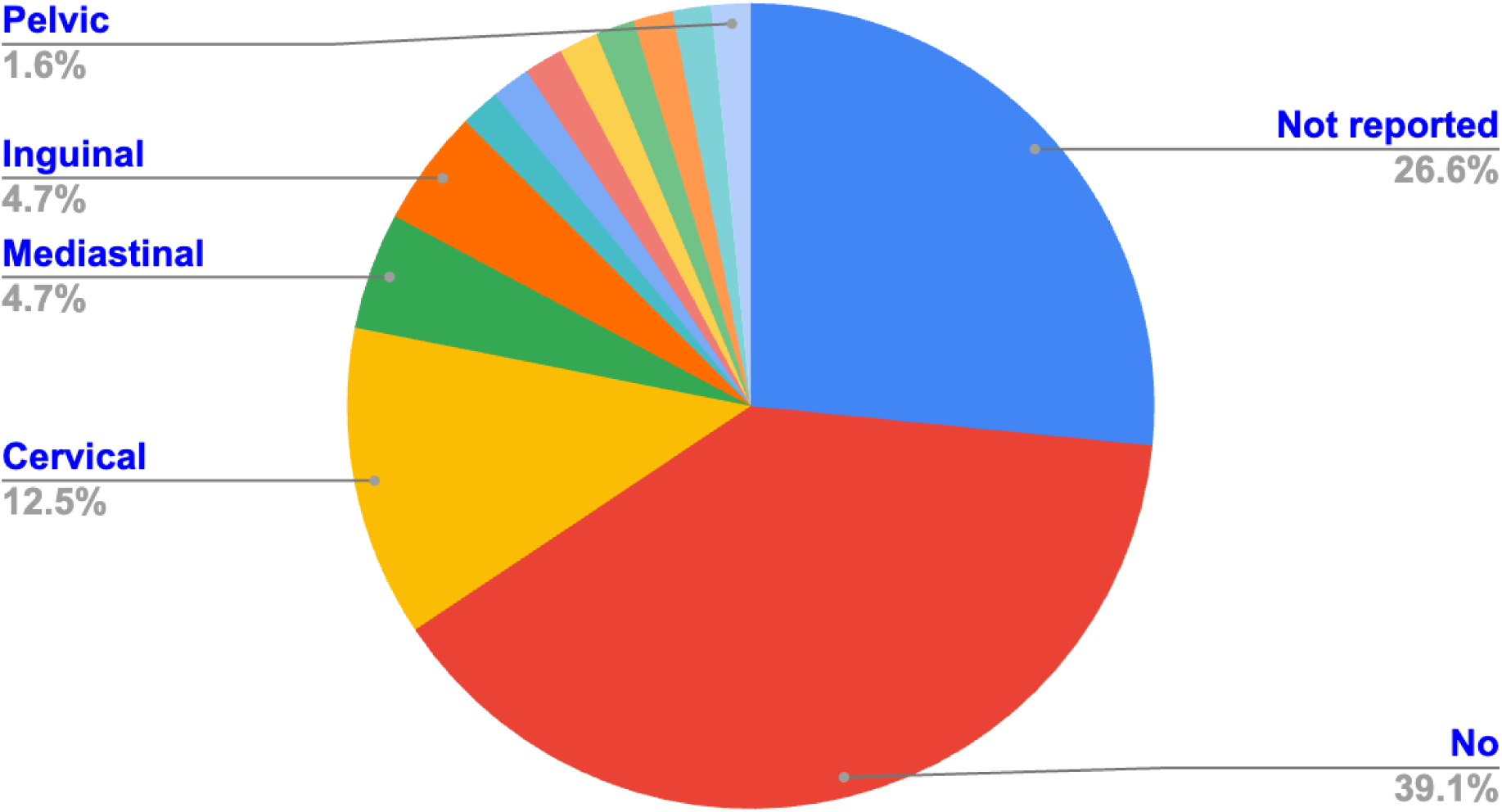
Distribution of Lymphadenopathy.

The top three sites of osseous involvement were Cranium, Tibia and Vertebrae (Fig. 3). Fifteen cases did not report extraosseous involvement whereas involvement of soft tissue was reported by 13 cases, lymph nodes by 8 cases, both lymph nodes and sinus by 3 cases, sinuses by 2 cases, both lymph nodes and soft tissue by 1 case and subcutaneous tissue by 1 case (Fig. 4). Out of the 53 cases studied, 10 displayed unusual symptoms^24,25,33,34,36,43,44,52,53^. Specifically, the symptoms related to the eyes were observed in 5 studies^24,33,34,43,53^, while ear-related symptoms were observed in 3 studies^24,34,36^ and paresthesia was seen only in 1 study^44^.

**Figure 3:**
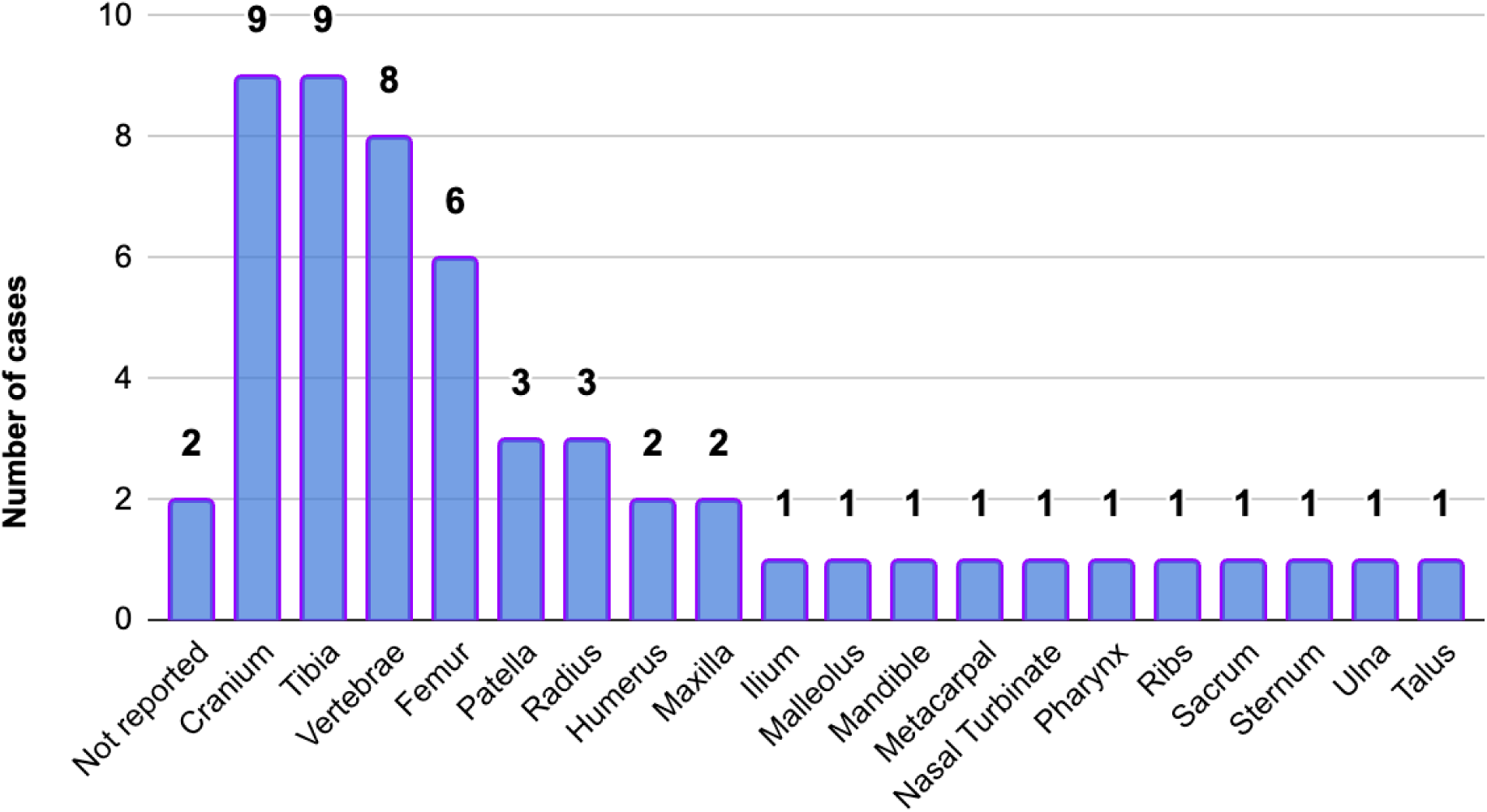
Osseous Involvement in RDD.

**Figure 4:**
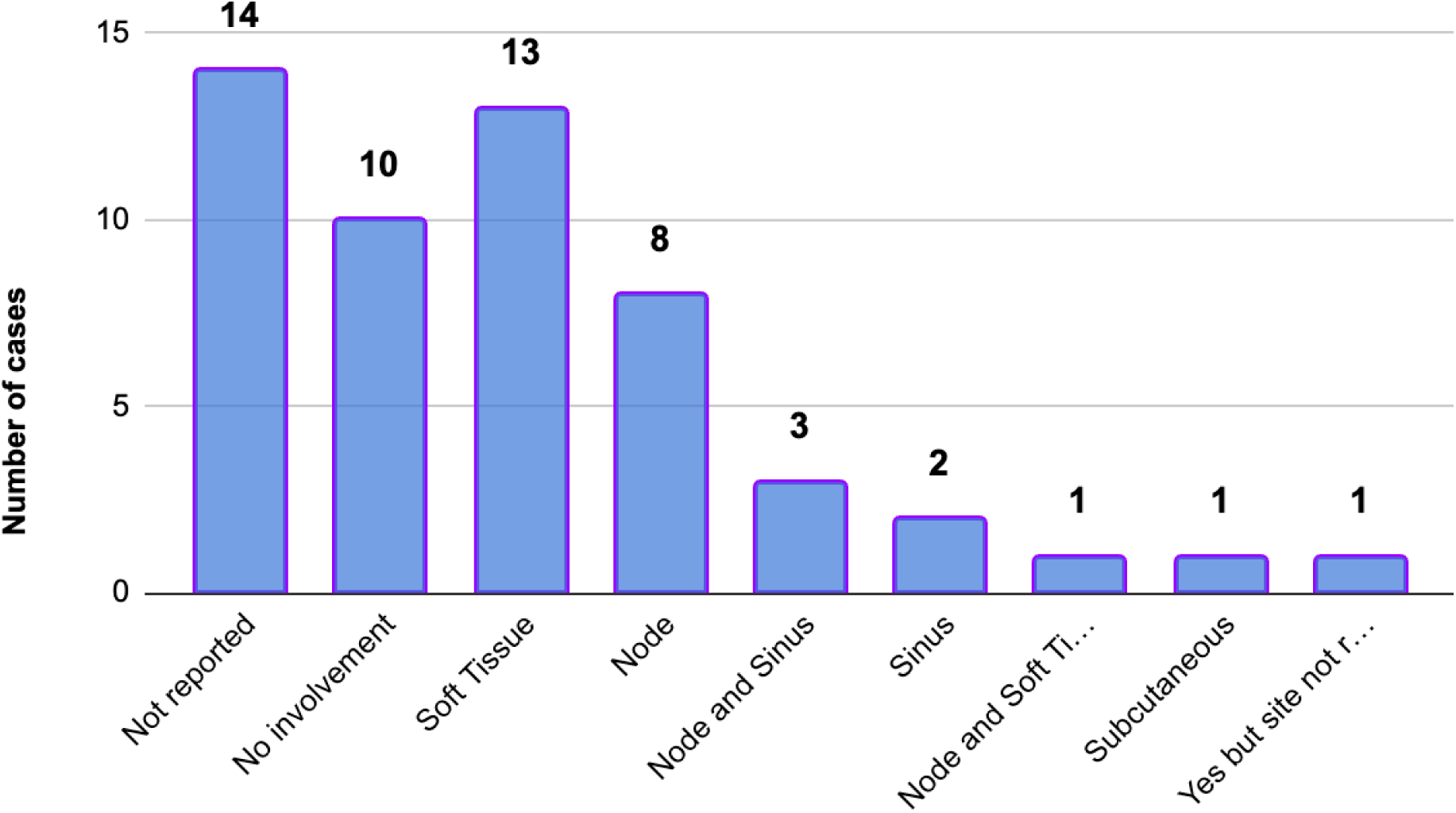
Extra-Osseous Involvement in RDD.

Regarding therapy, 41 cases received some form of treatment, 4 cases did not receive therapy, and 8 cases did not report their treatment status. The types of therapies administered included surgery in 30 cases, steroid treatment in 14 cases, chemotherapy in 5 cases, radiotherapy in 1 case, and immunosuppressants in 5 cases. Follow-up results indicated that 35 cases had a favorable outcome, 1 case had an unfavorable outcome, 2 cases were still under observation, and 15 cases did not report their follow-up results.

Regarding the immunohistochemical analysis, out of the 29 studies, 5^11,13,36,52,58^ showed a positive result for CD1a, while the rest were negative. Cyclin d1 levels were reported in only 2 studies, with Adam et. al.^57^ showing a positive result and Sciacca et. al.^44^ showing a negative result. Additionally, only in one study (Arun et. al.^21^) out of the 39 cases, S100 was negative.

Out of the total reported cases of RDD involving bone, a significant association was found between lymphadenopathy with steroid therapy (OR: 20.125, 95% CI: 3.02 to 134.09, P=0.0004) and surgery (P=0.1907). Extraosseous manifestation of RDD has also shown significant association with steroid therapy (P=0.0145), T1-isointense MRI (OR: 0.173, 95% CI: 0.0304 to 0.986, P=0.035), T1-hyperintense MRI (OR: 0.16, 95% CI: 0.0314 to 0.815, P=0.019) and T2-hyperintense MRI (OR: 0.115, 95% CI: 0.0206 to 0.6456, P=0.007). Odds ratio between extraosseous manifestation and steroid therapy could not be calculated.

## DISCUSSION

The relatively uncommon disorder RDD, described in 1969 by Rosai and Dorfman. Extranodal Rosai-Dorfman Disease (RDD) can affect various sites in the body, such as the skin, lymph nodes, bone, head and neck, kidney, CNS, breast, sinus, lung, liver, orbit, etc^7,8^. These sites are known as the preferred sites for the extra nodal form of RDD.

The study conducted by Mosheimer et al.^51^ revealed similar findings to our research. We observed that the cranium and long bones were the most commonly affected areas in cases of RDD. Additionally, in 29 out of 53 cases, there was evidence of extra osseous involvement, particularly in soft tissues, lymph nodes, and sinuses. Nasany RA et al.^60^ found that RDD involvement was most commonly observed in the dura, followed by the spine, brain parenchyma, orbits, and calvarium.

In contrast to our finding of lymphadenopathy in 20.75% of cases, a study on spinal RDD by Hu PP et al.^61^ reported lymph node enlargement in 17.4% cases, suggesting that spinal RDD might possess distinct pathological features. Furthermore, lymphadenopathy was a prevalent feature (64.5%) in pediatric RDD cases as reported by Alwani MM et al.^62^. Baeesa SS et al.^63^ revealed that RDD cells often accumulated in the lymph nodes, but they could also be found in extranodal sites such as the skin, soft tissue, upper respiratory tract, bones, eyes, retro orbital tissue, salivary glands, liver, pancreas, breast, and other locations.

Our study indicated that surgical intervention was performed in 30 out of 41 RDD cases, while steroids were administered in 14 out of 41 cases. Hu PP et al.^61^ recommended total resection as the preferred treatment option, particularly for isolated primary spinal RDD lesions. Surgical resection, either with or without adjuvant therapies, remained the primary approach when therapeutic interventions were necessary. Adjuvant therapies, such as steroids, chemotherapy, and radiotherapy, were reserved for progressive lesions and patients with systemic involvement. Nasany RA et al.^60^ suggested that chemotherapies had shown greater effectiveness in treating neurological Langerhans cell histiocytosis (LCH).

Like the findings of Alwani MM et al.^62^, our study demonstrated that surgical excision was the mainstay of treatment for RDD. Complete excision resulted in symptom reversal, low recurrence rates, and a positive long-term prognosis. Additionally, we observed that most patients with lymphadenopathy were treated solely with adjuvant therapy in addition to surgery, aligning with Alwani MM et al.^62^’s observations.

Immunohistochemistry plays a vital role in the diagnosis of RDD, as highlighted by Ojha J et al.^64^. S100 protein immunohistochemistry revealed strong cytoplasmic and nuclear reactivity in the histiocytes of RDD. These histiocytes were found to be positive for CD68, CD163, and Alpha-1-antitrypsin.

In our analysis we found that some cases present with unusual symptoms that delay their diagnosis. Similar observations were made by Ojha et al.^64^, further illustrating the need for accurate diagnosis and awareness of atypical manifestations of the disease. Osseous RDD involving bone may include other conditions that can cause bone lesions or masses, such as bone tumors (benign or malignant), histiocytic sarcoma (often characterized by marked cytologic atypia and frequent mitosis as well as necrosis), osteomyelitis, metastatic bone disease, and other non-neoplastic histiocytic disorders, such as, reactive histiocytosis (i.e. chronic osteomyelitis and infectious osteomyelitis with granulomatous inflammation), Langerhans cell histiocytosis and Erdheim-Chester disease. Differential diagnosis may also depend on the location and size of the bone lesion. It is essential for clinicians to conduct a thorough clinical evaluation, imaging studies, and biopsy to accurately diagnose RDD and distinguish it from other bone disorders.

Most of the literature included in this study consisted of case reports, which are considered to be low-quality evidence. This could potentially impact the overall quality of the study. There is a possibility of inclusion bias due to the search strategies employed and the limited availability of certain literature. There may also be a bias towards recognizing only symptomatic sites by the clinicians. Some of the included studies lacked complete records of the relevant data and reported information showed heterogeneity, which could potentially affect the statistical analysis and interpretation of the results.

In summary, RDD, a relatively uncommon disorder, affects diverse body sites, including skin, lymph nodes, bones, and various organs. Treatment involves surgical intervention and adjuvant therapies, with immunohistochemistry aiding diagnosis. However, atypical manifestations can lead to delayed recognition, underscoring the need for accurate diagnosis and awareness. RDD’s resemblance to other conditions necessitates thorough clinical evaluation and biopsy. Despite valuable insights into RDD characteristics and treatments from various studies, limitations such as case report dominance, biases in search strategies and data availability, and heterogeneity in reported information underscore the need for cautious interpretation of the findings.

## Author Contributions

Contributors:

Study design, literature review, statistical analysis: AA. Data management, data analysis, drafting manuscript: AA.

Access to data: AA, MA

Manuscript revision, intellectual revisions, mentorship: MA Final approval: AA, MA

## Acknowledgments

There were no further contributions to this project beyond those of the listed authors.

## Data Availability Statement

The original contributions presented in the study are included in the article/Supplementary Material, further inquiries can be directed to the corresponding author/s.

